# IL-6 trans-signaling mediates cytokine secretion and barrier dysfunction in hantavirus infected cells and correlate to severity in HFRS

**DOI:** 10.1101/2024.08.15.24312051

**Authors:** Kimia T. Maleki, Linda Niemetz, Wanda Christ, Julia Wigren Byström, Therese Thunberg, Clas Ahlm, Jonas Klingström

## Abstract

**Background:** Hantavirus causes hemorrhagic fever with renal syndrome (HFRS) and hantavirus pulmonary syndrome (HPS). Strong inflammatory responses and vascular leakage are important hallmarks of these often fatal diseases. The mechanism behind pathogenesis is unknown and no specific treatment is available. IL-6 was recently highlighted as a biomarker for HPS/HFRS severity. IL-6 signaling is complex and context dependent: while classical signaling generally provide protective responses, trans-signaling can cause severe pathogenic responses. This study aims to investigate a potential role for IL-6 trans-signaling in hantavirus pathogenesis.

**Methods:** Effects of IL-6 trans-signaling during *in vitro* hantavirus infection were assessed using primary human endothelial cells treated with recombinant soluble IL-6 receptor (sIL-6R). Plasma from Puumala orthohantavirus-infected HFRS patients (n=28) were analyzed for IL-6 trans-signaling potential and its associations to severity.

**Findings:** *In vitro*, sIL-6R treatment of infected cells enhanced IL-6 and CCL2 secretion, upregulated ICAM-1, and affected VE-cadherin leading to a disrupted cell barrier integrity. HFRS patients showed altered plasma levels of sIL-6R and soluble gp130 (sgp130) resulting in an increased sIL-6R/sgp130 ratio suggesting enhanced IL-6 trans-signaling potential. Plasma sgp130 levels negatively correlated with number of interventions and positively with albumin levels. Patients receiving oxygen treatment displayed a higher sIL-6R/sgp130 ratio compared to patients that did not.

**Interpretation:** IL-6 trans-signaling is linked to hantavirus pathogenesis. Targeting IL-6 trans-signaling might provide a therapeutic strategy for treatment of HPS and severe HFRS.

**Research in context:** *Evidence before this study:* IL-6 have dual effects, it provides important antiviral responses but can also cause severe pathogenesis. IL-6 mediated effects are activated by two different mechanisms, classical signaling and trans-signaling. IL-6 trans-signaling have recently been shown to be responsible for IL-6 mediated pathogenesis. Endothelial cells lack membrane-bound IL-6 receptor and only respond to IL-6 trans-signaling. Orthohantaviruses cause hemorrhagic fever with renal syndrome (HFRS) and hantavirus pulmonary syndrome (HPS; also called hantavirus cardiopulmonary syndrome (HCPS)), acute severe zoonotic diseases with high fatality rates for which specific treatments and vaccines are lacking. Deregulated vascular permeability and inflammation are hallmarks of HPS and severe HFRS. However, the mechanisms underlying hantavirus disease are unknown, hampering the development of treatments. Orthohantaviruses primarily infect endothelial cells. Previous studies have demonstrated a correlation between elevated IL-6 levels and increased severity of HPS and HFRS. Whether this link is causal, and if so the mechanisms behind it, has not been determined.

*Added value of this study:* This study demonstrates that hantavirus-infected endothelial cells produce large amounts of IL-6, and are highly sensitive to IL-6 trans-signaling mechanisms. Together this strongly amplifies the IL-6 production and disrupt the endothelial cell barrier causing severe fluid leakage. When analyzing a potential role for IL-6 trans-signaling in patients, we observed increased systemic IL-6 trans-signaling potential, and that this was linked to severity, in Puumala virus-infected HFRS patients.

*Implications of all the available evidence:* These data suggest the two hallmarks of HPS and HFRS; increased vascular permeability and strong inflammation, is linked via hantavirus-induced endothelial cell IL-6 production and IL-6 trans-signaling effects. Treatment targeting IL-6 trans-signaling might provide a therapeutic strategy for HPS and severe HFRS.

## INTRODUCTION

Orthohantavirus comprise a genus of single-stranded negative sense RNA viruses.^1^ Orthohantaviruses, hereafter referred to as hantaviruses, are transmitted to humans via inhalation of aerosolized virions from rodent excreta. Puumala orthohantavirus (PUUV) is the most prevalent hantavirus species in Europe and causes hemorrhagic fever with renal syndrome (HFRS).^2^ HFRS patients typically present with headache, fever, myalgia and renal symptoms.^2,3^ Fatality rates ranges between 0·4 to around 10%, depending on the specific hantavirus.^2,3^ In the Americas, hantaviruses such as Andes virus (ANDV) cause hantavirus pulmonary syndrome (HPS), a disease characterized by influenza-like symptoms and severe pulmonary dysfunction leading to a fatality rate of around 30-40%.^3^ Lung involvement is also seen in many PUUV-infected HFRS patients.^4^ Hantavirus primarily infects endothelial cells, especially in the lungs.^2^ Increased vascular permeability is a prominent hallmark of hantavirus infection, responsible for the life-threatening pulmonary dysfunction in HPS and severe HFRS.^2^ However, the mechanisms behind hantavirus-induced vascular permeability are not known. As hantavirus infected cells are protected from apoptosis,^5,6^ it is likely that other factors than endothelial cell death are involved in the etiology of vascular permeability during hantavirus infection.^2,3^

HFRS and HPS patients commonly exhibit increased systemic levels of pro-inflammatory cytokines.^7–11^ Recently, we and others reported that IL-6 is associated with increased disease severity in HPS.^7,12^ In addition, we showed that serum IL-6 levels are higher in fatal compared to non-fatal HPS cases.^7^ In PUUV-caused HFRS, high plasma IL-6 levels have been associated with high serum creatinine levels, thrombocytopenia and longer hospitalization, suggesting that IL-6 is associated also to the disease severity of HFRS.^13^ The mechanisms behind the association between IL-6 and disease severity of hantavirus infections are currently unknown.

IL-6 is a pro-inflammatory cytokine with many functions including in eliciting the acute phase response and promoting T cell activation and B cell maturation.^14^ IL-6 signals via a receptor complex constituting of the IL-6 receptor (IL-6R) and gp130.^14,15^ Classical IL-6 signaling is achieved upon binding of IL-6 to membrane bound IL-6R and gp130.^14,15^ While gp130 is expressed by all cells, IL-6R is expressed mainly by certain immune cells and hepatocytes. Thus, classical IL-6 signaling is restricted only to these IL-6R-expressing cell types.^14,15^ However, proteolytic cleavage and alternative splicing create soluble IL-6R (sIL-6R) which binds to IL-6 with low affinity.^16^ In turn, the soluble IL-6:sIL-6R complex can bind to gp130 on any cell and allow for so called IL-6 trans-signaling.^15,17^ Hence, trans-signaling allows for IL-6 signaling in cells with low or absent IL-6R expression. Recently, it has become evident that trans-signaling is the predominant pathway behind IL-6 mediated pathogenesis.^18^ Also gp130 exists as a soluble version, soluble gp130 (sgp130), produced by proteolytic cleavage as well as alternative splicing.^19^ sgp130 can bind to the IL-6:sIL-6R complex, hindering binding to membrane bound gp130, thereby inhibiting trans-signaling.^19^ In blood, sIL-6 and sgp130 levels are high, and their ratio will determine the IL-6 trans-signaling potential, thereby affecting the potential pathogenic effects of IL-6 in circulation.^18^

Endothelial cells can produce high levels of IL-6 upon stimulation.^20^ As endothelial cells express no or very little IL-6R, they *per se* do not seem to respond to the IL-6 they produce.^20–23^ However, studies have shown that addition of sIL-6R to endothelial cell cultures renders them responsive to IL-6 via trans-signaling.^21^

Here, we sought to investigate the source of IL-6 during hantavirus infection, as well as the possible consequences of IL-6 trans-signaling on infected vascular endothelial cells. We show that endothelial cells produce large amounts of IL-6 upon PUUV infection. Addition of sIL-6R to PUUV infected endothelial cells lead to strongly enhanced secretion of IL-6 and CCL2 and upregulation of ICAM-1 on the cell surface. In addition, we show that sIL-6R led to VE-cadherin internalization and increased monolayer permeability in infected endothelial cells. Finally, we show that HFRS patients exhibit altered levels of sIL-6R and sgp130, and that this correlate to markers of severity, suggesting a direct role for IL-6, via IL-6 trans-signaling, in hantavirus pathogenesis.

## METHODS

### Patients

Twenty-eight HFRS patients were included in the study. Patients were diagnosed during the years 2006-2014, at the University Hospital of Umeå, Sweden. Twenty uninfected controls sampled in 2017 were included as control subjects. Peripheral blood was collected into CPT tubes, as described previously.^24^ Following centrifugation, plasma was retrieved and stored at −80°C until analysis. All subjects provided written consent before participation in the study. Ethical approval was obtained from the Regional Ethics Committee of Umeå University (application number 04-133M).

The HFRS cohort included 13 females and 15 males with a mean age of 49 years (range 18-78 years) and the controls included 7 females and 13 males of a mean age of 50 years (range 37-63 years) (Table 1). Samples were collected from the acute phase at a median of 5 days (range 2-7 days) post symptom debut, and from the convalescent phase at a median of 98 days (range 42-494 days) post symptom debut (Table 1).

**Table 1.** Patient characteristics and clinical data.

| Parameter | Controls | HFRS patients |  |
| --- | --- | --- | --- |
|  |  | Acute phase | Conv. phase |
| No. of patients | 20 | 28 |  |
| Gender (female/male) | 7/13 | 13/15 |  |
| Age (years), mean (range) | 50 (37-63) | 49 (18-78) |  |
| Days post symptoms debut, median (range) | NA | 5 (2-7) | 63 (42-494) |
| WBC count ($\times 10^9/L$), mean ± SD* | n.d. | $8.9 \pm 5.3$ | $7.1 \pm 2.5$ |
| Platelet count ($\times 10^9/L$), mean ± SD<sup>#</sup> | n.d. | $91 \pm 49$ | $276 \pm 46$ |
| Serum creatinine ($\mu\text{mol/L}$), mean ± SD<sup>§</sup> | n.d. | $204 \pm 150$ | $87 \pm 41$ |
| CRP (mg/L), mean ± SD<sup>€</sup> | n.d. | $75 \pm 47^a$ | $4 \pm 3^b$ |
| Serum albumin (g/L), mean ± SD<sup>†</sup> | n.d. | $32 \pm 5.8^c$ | $42 \pm 3.5^d$ |
| Intravenous fluid, no. of patients (%) | NA | 19/28 (68) | NA |
| Platelet transfusion, no. of patients (%) | NA | 3/28 (10) | NA |
| Oxygen treatment, no. of patients (%) | NA | 8/28 (29) | NA |
| Severe bleeding, no. of patients (%) | NA | 4/28 (14) | NA |
| Thrombosis, no. of patients (%) | NA | 4/28 (14) | NA |

Abbreviations: HFRS, hemorrhagic fever with renal syndrome; NA, not applicable; WBC, white blood cell; n.d., not done; CRP, C-reactive protein.
\*WBC; normal range, $3.5\text{--}8.8 \times 10^9/L$ .
<sup>#</sup>Platelet count; normal range, $165\text{--}387 \times 10^9/L$ for women, $145\text{--}348 \times 10^9/L$ for men.
<sup>§</sup>Serum creatinine; reference value $<90 \mu\text{mol/L}$ for women, $<105 \mu\text{mol/L}$ for men.
<sup>€</sup>CRP; reference value $<3 \text{ mg/L}$ .
<sup>†</sup>Serum albumin; normal range $34\text{--}54 \text{ g/L}$ .
<sup>a</sup> $n=26$ .
<sup>b</sup> $n=25$ .
<sup>c</sup> $n=13$ .
<sup>d</sup> $n=9$ .

### Cells and viruses

Pooled human umbilical vein endothelial cells (HUVECs) (Lonza) were maintained in endothelial growth medium (EGM-2) supplemented with EGM-2 endothelial SingleQuots (Lonza) in 5% CO_2_ at 37°C. Prior to experiments, hydrocortisone was excluded from the medium. Buffy coats from blood donors were purchased from Karolinska University Hospital (Stockholm, Sweden). Peripheral blood mononuclear cells (PBMCs) were isolated from buffy coats using Lymphoprep (Stemcell Technologies) and maintained in RPMI-1640 medium (GE Healthcare) supplemented with 10% FCS (Sigma-Aldrich) and 2 mM L-glutamine (Life Technologies). PUUV strain CG1820 was propagated on A549 cells and ANDV strain Chile-9717869 on Vero E6 cells, as previously described.^25^

### Infection and treatments

HUVECs were infected with 200 µl (in 24 well plates) or 1 ml (in 6 well plates) virus diluted in HUVEC medium at multiplicity of infection (MOI) 1. Cells were infected for 1 h in 5% CO_2_ at 37°C, with gentle shaking every 10 min. After infection, virus was removed and replaced with 1 ml fresh medium. At 48 h post infection, medium was replaced with medium containing recombinant human IL-6R alpha protein (R&D systems) at different concentrations. Cells without sIL-6R treatment and cells treated with 10 ng/ml recombinant IL-6 (rIL-6) (R&D systems), in addition to the sIL-6R treatment, were used as controls. After 24 h treatment, supernatants were collected and stored at −80°C until analysis.

PBMCs cultured in 96-well plates (1 million cells in 200 µl medium) were exposed to PUUV (MOI=3) for 2 h in 5% CO_2_ at 37°C. After infection, virus was removed and replaced with fresh medium. Supernatants were collected after a centrifugation step and stored at −80°C until analysis.

### Flow cytometric analyses

HUVECs were detached using Accutase (Thermo Fisher Scientific) and then used for flow cytometry. HUVECs were stained with anti-ICAM-1 antibody conjugated with PE-Vio770 (Milteny Biotec) for 20 min at room temperature (RT). Live/Dead Aqua (Invitrogen) was used for the identification of dead cells. Cells were fixed for 30 min using Transcription Factor Staining Buffer Set (BD Biosciences). Samples were acquired on a BD LSR Fortessa instrument (BD Biosciences). Data were analyzed using FlowJo version 10.4.

### ELISA

Prior to ELISA, plasma samples were diluted in ready-to-use ELISA diluent (Mabtech); 1:2 for IL-6 and IL-6/sIL-6R complex and 1:400 for sIL-6R and sgp130 ELISAs. Levels of IL-6 were analyzed using ELISA development kit (Mabtech) and levels of sIL-6R, IL-6:IL-6R complex, and sgp130 were analyzed using DuoSet ELISA kits (R&D), all according to the manufacturer’s guidelines.

### Immunofluorescence

HUVECs cultured on glass cover slips were fixed with pre-warmed 4% paraformaldehyde for 15 min at RT. Cells were then permeabilized using 0·5% Triton X for 5 min at RT, washed three times and blocked with 0·5% BSA in PBS for 30 min at RT. Cover slips were incubated with primary antibody for 1 h at RT, washed in PBS three times and then incubated with secondary antibody for 1 h at RT. VE-cadherin expression was detected using anti-VE-cadherin monoclonal antibody (Cell Signaling Technology) and goat anti-rabbit IgG AF488 (Life Technologies). PUUV proteins were detected using polyclonal antibodies from convalescent patient serum for 1 h at RT and goat anti-human IgG AF647 (Life Technologies). Nuclei were stained using DAPI (Life Technologies). Washed cover slips were mounted onto glass slides using ProLong Gold Antifade Mountant (Thermo Fisher Scientific). Cells were examined by immunofluorescence confocal microscopy at 60X magnification. Images were analyzed using ImageJ.

### Transendothelial electrical resistance (TEER)

HUVECs, uninfected or infected with PUUV for 24 h, were split onto HTS Transwell 24-well plates with 0·4 μm pore and 6·5 mm inserts (Corning) at a density of 1×10^5^ cells in 100 µl per well. To the lower compartment, 600 µl medium was added. Cells were cultured at 37°C for 24 h and then treated with sIL-6R. The transendothelial electrical resistance (TEER) was measured after 24 h using an EVOM2 epithelial voltohmmeter (World precision instruments), according to the manufacturer’s guidelines. The mean TEER of three measurements/well was used to calculate the TEER/cm2.

### Statistical analyses

Statistical analyses were performed using Graph Pad Prism v.9. Paired comparisons within HFRS patients were performed using Wilcoxon test. Comparisons between controls and acute and convalescent HFRS patients were performed using Kruskal-Wallis test followed by Dunn’s multiple comparison test. Comparisons between *in vitro* conditions were performed using two-way ANOVA followed by Dunnet’s or Šídák’s multiple comparison test. Spearman’s rank correlation coefficient was used for examining correlations.

## RESULTS

### PUUV-infected cells demonstrate potent IL-6 secretion

To investigate possible sources of IL-6 production during hantavirus infection, we assessed IL-6 secretion from HUVECs and PBMCs. HUVECs were infected with PUUV at MOI 1 and supernatants were collected at 24, 48, and 72 h post infection. Infected HUVECs produced higher levels of IL-6 than uninfected HUVECs at 48 and 72 h post infection (Figure 1A). PBMCs were exposed to PUUV at MOI 3 and supernatants were collected after 24, 48, and 72 h. At 48 h post PUUV-exposure, PBMCs produced higher levels of IL-6 compared to unexposed PBMCs, albeit at lower concentrations than HUVECs (Figure 1B). Next, we examined the concentrations of sgp130 in the supernatants. In infected HUVECs, the levels of sgp130 were significantly increased at 72 h post infection (Figure 1C). In supernatants of PUUV-exposed PBMCs, no such increase was observed (Figure 1D). In the PBMC supernatants, also sIL-6R levels were analyzed. No significant increase in sIL-6R levels was observed in PUUV-exposed compared to unexposed PBMCs (Figure 1D). Together, these data show that PUUV induces strong IL-6 secretion in HUVECs and PBMCs, and that this coincides with increased production of sgp130 in HUVECs.

**Fig. 1.**
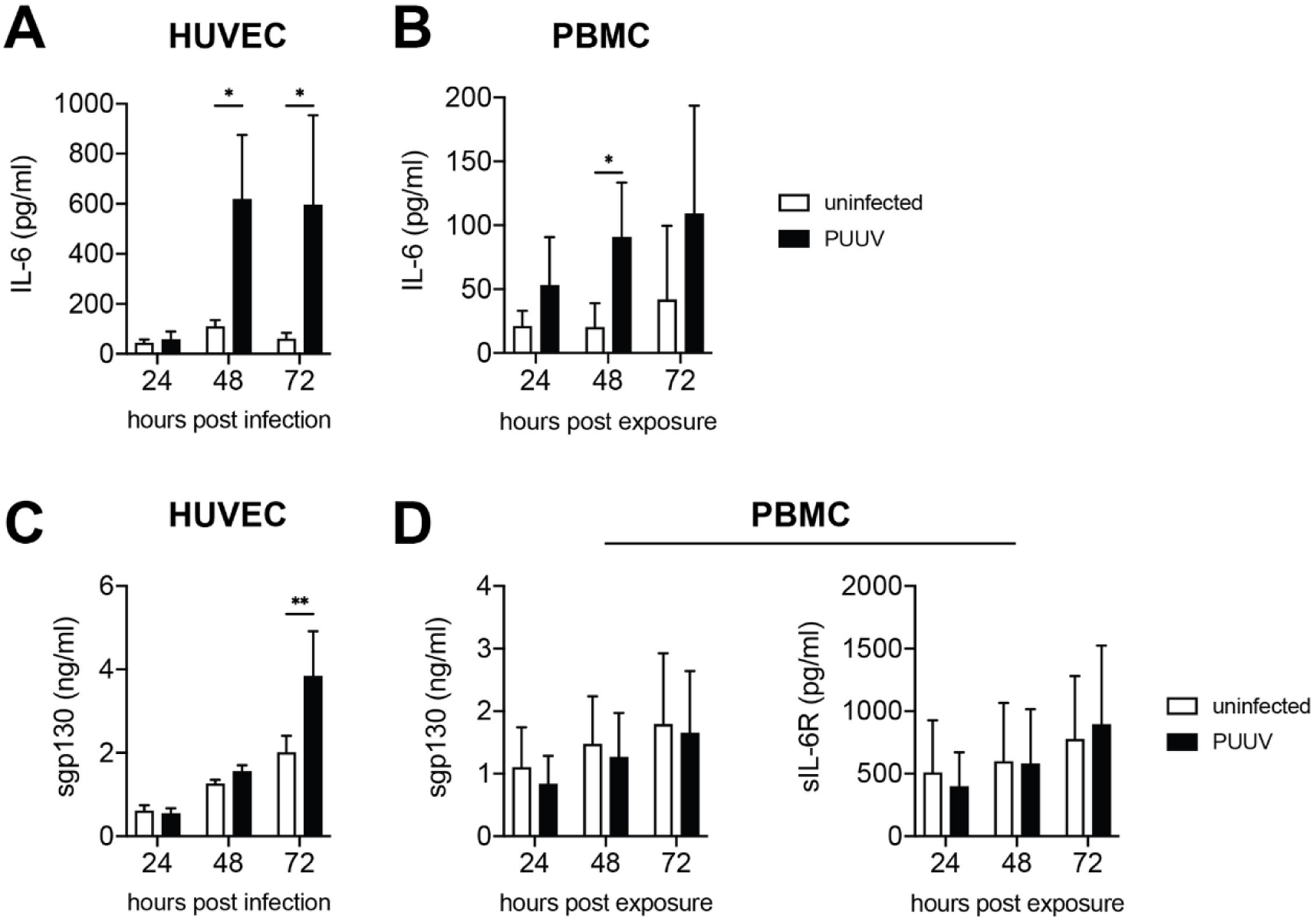
PUUV-infected cells secrete IL-6. HUVECs were infected with PUUV (MOI=1) and PBMCs were exposed to PUUV (MOI=3) for 24-72 h. Supernatants were assessed for IL-6, soluble gp130 (sgp130), and soluble IL-6R (sIL-6R) using ELISA. (**A**) Levels of IL-6 in supernatants of HUVECs (n=3) and (**B**) PBMCs (24-48 h, n=6, 72 h, n=2). (**C**) sgp130 levels in supernatants of HUVECs (n=3). (**D**) Levels of sgp130 (n=2) and sIL-6R (n=7) in PBMC supernatants. Two-way ANOVA followed by Šídák’s multiple comparison test. *, p<0·05, **, p<0·01.

### IL-6 trans-signaling in PUUV-infected cells drives inflammation

Having shown that HUVECs produce high levels of IL-6 upon PUUV infection, we next investigated potential autocrine effects of IL-6 trans-signaling on endothelial cells. IL-6 trans-signaling in endothelial cells has been shown to increase secretion of IL-6,^21^ and CCL2.^22,24^ As endothelial cells have been reported to be unresponsive to IL-6 treatment,^21^ inflammatory responses downstream of IL-6 were assessed after addition of recombinant sIL-6R to the cell cultures. At 48 h post infection, HUVEC medium was exchanged to fresh medium with or without different concentrations of sIL-6R. After 24 h of treatment, IL-6 and CCL2 levels in supernatants were determined by ELISA. Levels of IL-6 and CCL2 in supernatants were increased in infected HUVECs treated with sIL-6R, in a dose-dependent manner (Figure 2A-B). As expected,^22,26^ increased CCL2 secretion was also observed in uninfected HUVECs treated with recombinant IL-6 (rIL-6) along with the sIL-6R treatment (Figure S1A).

**Fig. 2.**
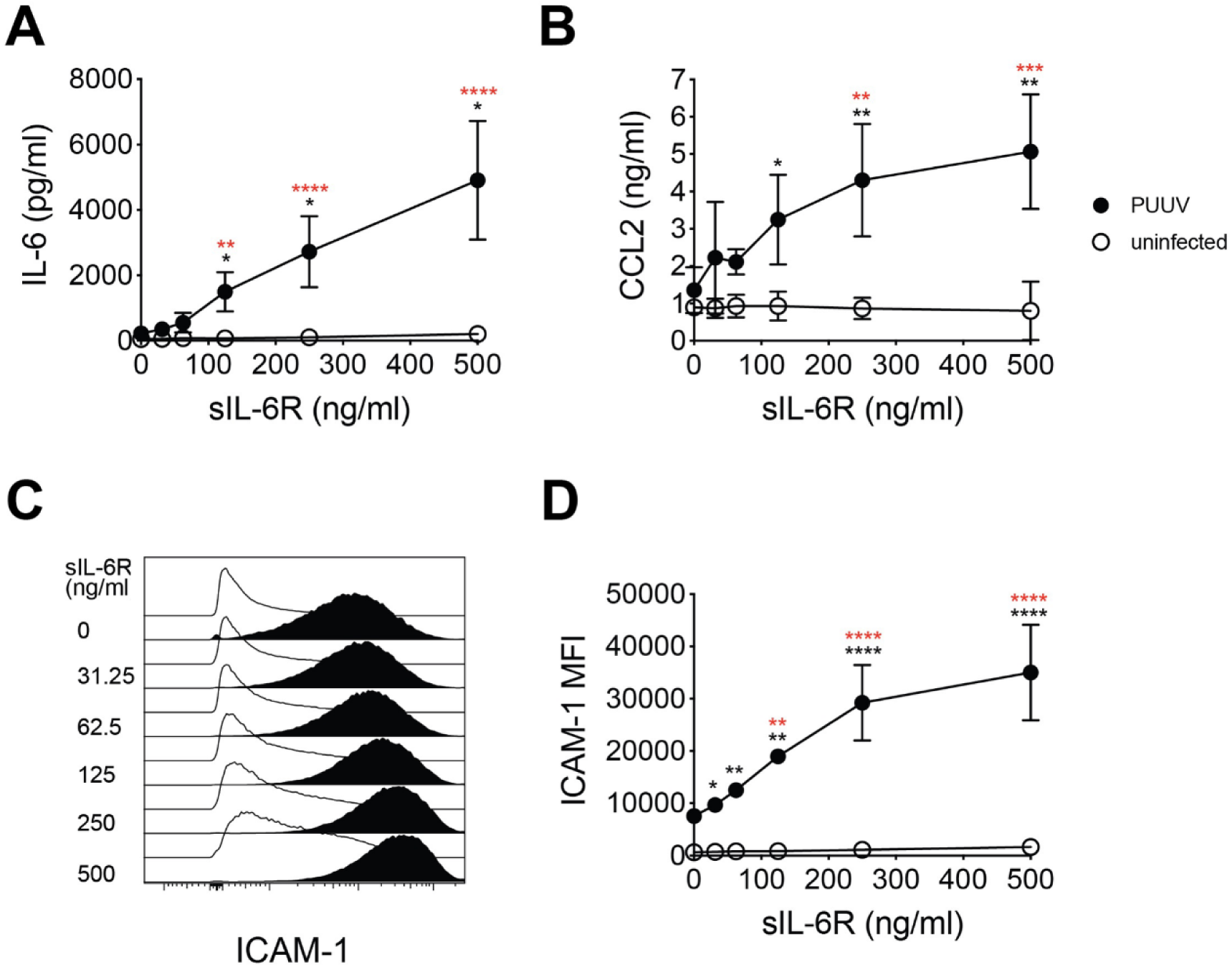
IL-6 trans-signaling activates endothelial cells and drives inflammation. HUVECs were infected with PUUV (MOI=1) for 48 h and then treated with sIL-6R at the concentrations 31·25, 62·5, 125, 250, or 500 ng/ml for 24 h or left untreated. (**A**) Levels of IL-6 (n=5) and (**B**) CCL2 (n=3) in supernatants of uninfected and PUUV-infected HUVECs. (**C**) Representative histogram plot and (**D**) graph showing median ICAM-1 expression on infected and uninfected HUVECs (n=3). Symbols depict mean and error bars indicate SD. Two-way ANOVA followed by Dunnet’s or Šídák’s multiple comparison test. Black asterisks indicate significance when comparing PUUV to uninfected. Red asterisks indicate significance when comparing each sIL-6R-treated conditions of PUUV-infected cells with untreated PUUV-infected cells. *, p<0·05; **, p<0·01; ***, p<0·001, ****, p<0·0001.

IL-6 signaling has been reported to increase the cell surface expression of ICAM-1 on endothelial cells.^26–29^ Thus, ICAM-1 expression on HUVECs was assessed by flow cytometry. As previously observed for Hantaan orthohantavirus (HTNV) infected cells,^30,31^ increased ICAM-1 expression was observed on PUUV-infected compared to uninfected HUVECs (Figure 2C-D). Addition of sIL-6R to the cultures further increased the ICAM-1 expression on infected, but not uninfected cells, in a dose-dependent manner (Figure 2C-D). Together, these data indicate that endogenously produced IL-6 from PUUV-infected endothelial cells, in presence of sIL-6R, stimulate IL-6 trans-signaling in an autocrine manner that fuels the inflammatory responses by causing endothelial cell activation and increased secretion of IL-6 and CCL2.

Levels of IL-6 in supernatants were also increased in ANDV-infected HUVECs treated with sIL-6R (Figure S2), suggesting this is a common feature of HFRS and HPS-causing hantaviruses.

### PUUV-mediated IL-6 trans-signaling disrupts endothelial cell barrier functions

To further investigate the functional consequences of IL-6 trans-signaling, we sought to evaluate the barrier function in infected HUVEC monolayers. For this purpose, we first examined the expression of VE-cadherin in infected and uninfected cells, with or without sIL-6R, using immunofluorescence microscopy. In uninfected untreated cells, solid VE-cadherin junctions and intact cell monolayers were observed (Figure 3A). Addition of sIL-6R to uninfected cells had no clear effect on the VE-cadherin organization nor the cell integrity (Figure 3A). In contrast, internalization of VE-cadherin was seen in PUUV-infected HUVECs without sIL-6R treatment (Figure 3A). sIL-6R treatment of infected cells caused further downmodulation of VE-cadherin from the cell surface (Figure 3A). Interestingly, while the cell monolayer appeared to be intact in untreated PUUV-infected cells, sIL-6R treatment caused gap formation in the cell monolayer, indicating that the barrier function was severely disrupted (Figure 3A). As expected, a similar phenotype was observed when uninfected cells were treated with rIL-6 in addition to the sIL-6R treatment (Figure S1B), showing that this indeed was dependent on IL-6 trans-signaling.

**Fig. 3.**
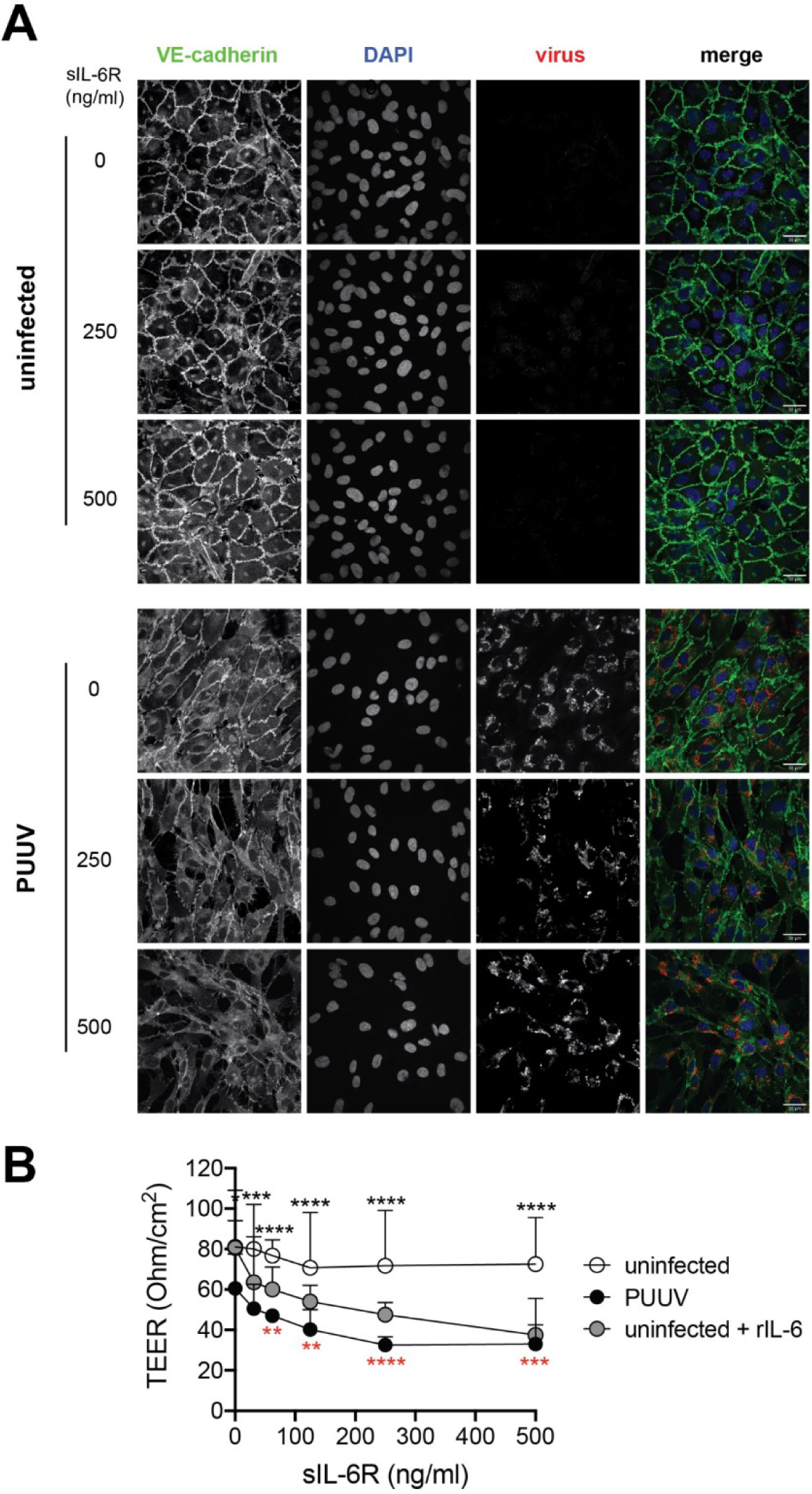
IL-6 trans-signaling disrupts endothelial cell barrier functions during hantavirus infection. Uninfected and infected HUVECs were treated with sIL-6R at the concentrations 31·25, 62·5, 125, 250, or 500 ng/ml for 24 h or left untreated. (**A**) Immunofluorescence images showing expression of DAPI (blue), virus (red), and VE-cadherin (green). Representative images of three independent experiments are shown. (**B**) Transendothelial electrical resistance of uninfected (white symbol) and infected HUVECs (black symbol), with and without sIL-6R. Uninfected HUVECs treated with rIL-6 in addition to sIL-6R were used as control (grey symbol). Symbols depict mean and error bars indicate SD. Two-way ANOVA followed by Dunnet’s or Šídák’s multiple comparison test. Black asterisks indicate significance when comparing PUUV to uninfected. Red asterisks indicate significance when comparing each sIL-6R-treated conditions of PUUV-infected cells with untreated PUUV-infected cells. *, p<0·05; **, p<0·01; ***, p<0·001, ****, p<0·0001.

To further examine the endothelial cell barrier function upon sIL-6R treatment, we analyzed the transendothelial electrical resistance (TEER) in PUUV-infected and uninfected HUVECs. PUUV infection alone caused decreased TEER compared to uninfected cells (Figure 3B). PUUV infection together with sIL-6R treatment further caused a dose-dependent loss of barrier function 24 h post treatment, as indicated by decreased TEER compared to uninfected HUVECs and PUUV-infected untreated cells (Figure 3B). Similarly, a dose-dependent decrease in TEER was observed when uninfected cells were treated with rIL-6 + sIL-6R (Figure 3B). Supporting previous reports,^32,33^ rIL-6 treatment alone did not affect the monolayer permeability in uninfected HUVECs (Figure 3B). Collectively, these data indicate that IL-6 produced by PUUV-infected endothelial cells in an IL-6 trans-signaling dependent manner enhances VE-cadherin disorganization and loss of endothelial cell monolayer integrity.

### The plasma sIL-6R/gp130 ratio is increased during acute HFRS

Given the pronounced effects of IL-6 trans-signaling observed in hantavirus infected cells *in vitro*, we next sought to evaluate the levels of soluble IL-6 receptors in HFRS patients. The physiological effects of IL-6 largely depend on the levels of sIL-6R and sgp130 in circulation.^15,18^ Although increased systemic IL-6 levels have been repeatedly reported in HFRS and HPS patients,^7–9,11,12^ the concentrations of sIL-6R and sgp130 have not yet been comprehensively studied in hantavirus-infected patients. Thus, we analyzed the levels of IL-6, sIL-6R, IL-6:sIL-6R complex, and sgp130 in plasma of 28 PUUV-infected HFRS patients, during acute and convalescent phase, as well as in 20 uninfected controls. The HFRS patients displayed typical symptoms for HFRS, showing thrombocytopenia, and elevated levels of creatinine and CRP (Table 1). Information regarding complications (thrombosis, severe bleeding) as well as information on medical interventions including administration of intravenous fluid, oxygen treatment, and platelet transfusion are presented in Table 1. IL-6 levels were, as previously reported,^8,11^ increased in acute HFRS (Figure 4A). Levels of sIL-6R were higher during acute, compared to the convalescent, HFRS, and similar to that observed in uninfected controls (Figure 4B). Levels of the IL-6:sIL-6R complex were not significantly altered during HFRS (Figure 4C) while sgp130 levels were decreased in both acute and convalescent HFRS, compared to controls (Figure 4D). sgp130 binds to the IL-6:sIL-6R complex thereby inhibiting binding of the complex to membrane bound gp130.^17^ Thus, when assessing the likelihood of IL-6 trans-signaling, the proportion of sIL-6R in relation to sgp130 is important to consider. To better assess the relation of the IL-6 receptors during HFRS, the ratio between sIL-6R and sgp130 was calculated. Interestingly, the sIL-6R/sgp130 ratio was significantly increased during acute HFRS, as compared to convalescent HFRS and controls (Figure 4E). Taken together, this suggests increased IL-6-trans-signaling potential during acute HFRS.

**Fig. 4.**
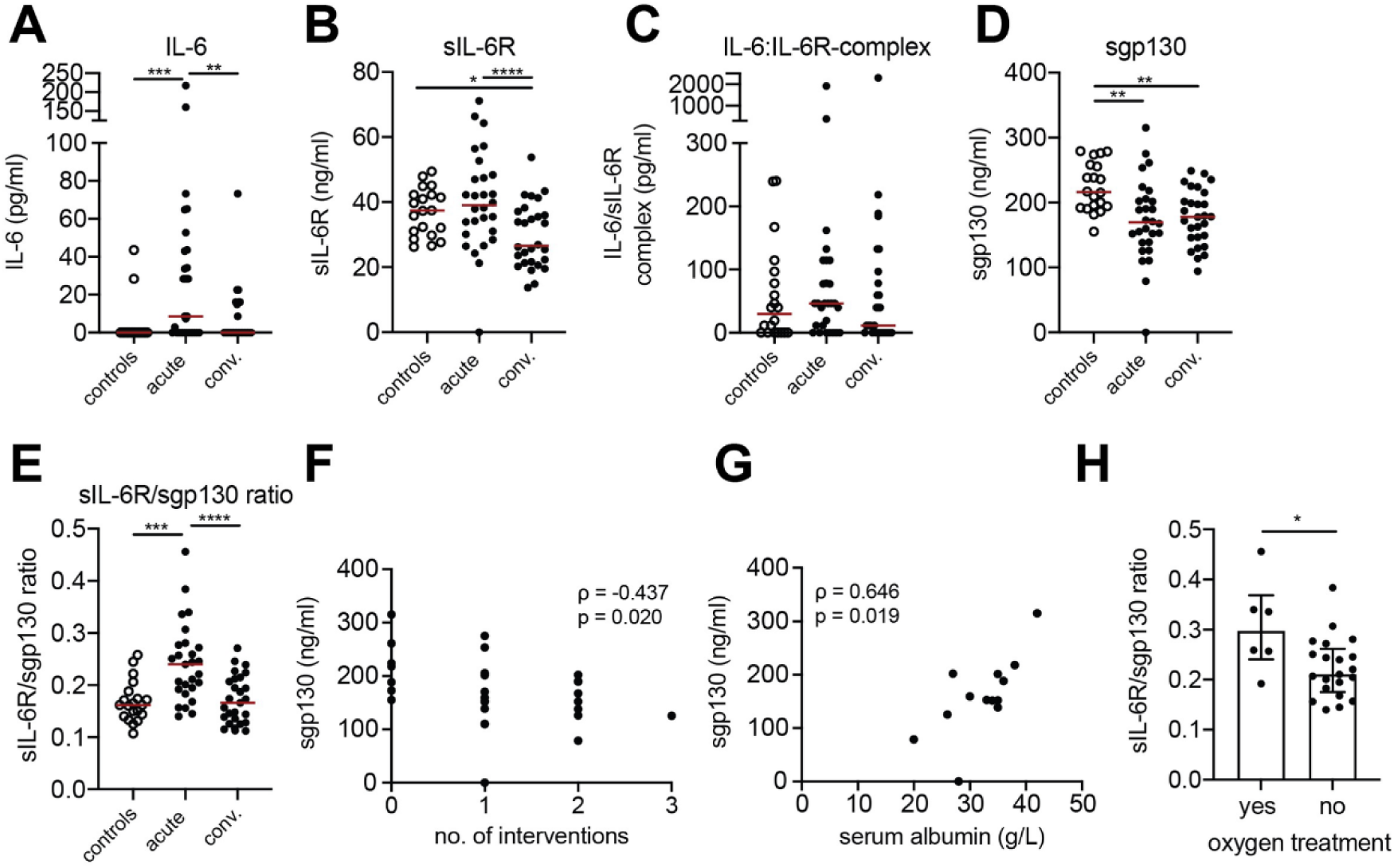
The plasma sIL-6R/sgp130 ratio is increased during acute HFRS and IL-6 trans-signaling potential correlate to severity. Plasma levels of (**A**) IL-6, (**B**) sIL-6R, (**C**) IL-6:sIL-6R complex, and (**D**) sgp130 in controls (n=20) and acute and convalescent HFRS patients (n=28). (**E**) Ratio of plasma sIL-6R and sgp130 in controls and HFRS patients (n=27). (**F**) Correlation between sgp130 levels and number of interventions during acute HFRS (n=28). (**G**) Correlation between sgp130 levels and serum albumin (n=13). (**H**) Plasma sIL-6R/sgp130 ratio in patients with or without oxygen treatment (median, interquartile range). Wilcoxon test; Kruskal-Wallis test; median. Spearman’s rank correlation coefficient. *, p<0·05; **, p<0·01; ***, p<0·001, ****, p<0·0001.

### Sgp130 level and IL-6 trans-signaling potential correlate to disease severity

We finally analyzed for possible correlations of IL-6 signaling factors, including IL-6 trans-signaling potential, and severity of HFRS. This revealed that sgp130 levels were inversely correlated with the number of interventions in patients (Figure 4F), suggesting that patients with a more complicated disease had the lowest sgp130 levels. Furthermore, sgp130 levels positively correlated with serum albumin levels (Figure 4G), indicating that decreased sgp130 levels coincide with loss of serum albumin, which is indicative of increased vascular permeability. The sIL-6R/sgp130 ratio was significantly higher in patients receiving oxygen treatment (Figure 4H). Together, this suggest low sgp130 levels and high IL-6 trans-signaling potential is associated with severe HFRS.

## DISCUSSION

HFRS and HPS are characterized by strong inflammatory responses and vascular permeability.^2,3,7–11^ Despite significant morbidity and mortality in hantavirus-infected individuals, no specific treatments are available. Importantly, the mechanisms driving hantavirus-mediated pathogenesis are not known, hampering development of specific therapeutics. IL-6 has been highlighted as an important cytokine in hantavirus infections, as it is associated with HFRS and HPS disease severity.^7,12,13^ However, the potential role for IL-6 in the pathophysiology of hantavirus diseases is unknown. Here, we report IL-6 trans-signaling as a potential mechanism behind IL-6-mediated pathology during hantavirus infections.

We observed that PUUV infection stimulates IL-6 secretion in HUVECs and PBMCs. IL-6 secretion has previously been reported in cells infected with ANDV, HTNV, and Prospect Hill orthohantavirus.^34,35^ When studying IL-6 signaling *in vitro*, it is important to consider the receptor availability on the cells of the model system. While gp130 is expressed by all cells, IL-6R is mainly expressed by hepatocytes and certain immune cells.^14^ Endothelial cells express no or very little IL-6R, and therefore display no or limited responses to IL-6 *per se*.^20,21,36^ This may explain why a previous study did not observe any effect upon IL-6 treatment of hantavirus infected cells.^32^ However, treatment of endothelial cells with IL-6 in combination with sIL-6R leads to IL-6 trans-signaling. IL-6 trans-signaling in endothelial cells has been shown to cause secretion of IL-6 and CCL2 as well as upregulation of ICAM-1 on the cell surface.^21,22,26–29^ Here, we recapitulated these findings in a PUUV-infection model. Using this model, we showed that PUUV-induced endogenous IL-6, in presence of sIL-6R, activated endothelial cells in an autocrine manner. This activation caused upregulation of the adhesion molecule ICAM-1, indicating enhanced activation of the endothelial cells. This is in line with the increased plasma levels of soluble ICAM-1, VCAM-1, E-selectin, and syndecan-1 observed in HFRS patients.^37^ PUUV-mediated IL-6 trans-signaling further promoted a pro-inflammatory loop with augmented secretion of IL-6 and CCL2. CCL2 is a known chemoattractant for T cells and myeloid cells.^38,39^ Apart from its chemoattracting function, CCL2 also mediates attachment to vascular endothelial cells and trans-endothelial migration of T cells and myeloid cells.^38–40^ Together, these data suggest that infected endothelial cells may, via IL-6 trans-signaling, constitute important drivers of IL-6 production and inflammation in HFRS/HPS patients.

Vascular permeability is a common hallmark of hantavirus infections.^2,3^ The mechanism behind why hantavirus infection leads to massively increased vascular permeability, including the life-threatening pulmonary dysfunction observed in HPS, is largely unknown.^3^ Here, we demonstrated that sIL-6R treatment of infected endothelial cells increases VE-cadherin disorganization and barrier permeability. This is in line with previous reports showing VE-cadherin internalization and increased permeability of endothelial cells treated with IL-6 in combination with sIL-6R.^28,33,41^ Remarkably, PUUV infection alone, without sIL-6R treatment, caused some VE-cadherin internalization and decreased monolayer integrity. Previously, a VEGF/VEGFR2-dependent internalization of VE-cadherin has been described in ANDV and HTNV infected endothelial cells.^42,43^ Such a VEGF-dependent mechanism could possibly explain the VE-cadherin disorganization and decreased membrane integrity seen in untreated PUUV-infected HUVECs. Taken together, these findings indicate that hantavirus infection alone to some extent modulates the endothelial cell barrier integrity, and that subsequent IL-6 trans-signaling further aggravates this, leading to a severe loss of barrier function.

The *in vivo* effects of IL-6 depend on the complex interactions of IL-6 with sIL-6R and sgp130, which constitute a buffer system that regulates the half-life and signaling of IL-6.^15,18^ While it is well-established that IL-6 levels are increased in HFRS and HPS patients, peripheral levels of sIL-6R and sgp130 have not been extensively studied in hantavirus infected patients. Here, plasma sIL-6R levels were found to be elevated during the acute, compared to the convalescent, phase of HFRS. Intriguingly, compared to controls, lower plasma sgp130 concentration was observed both during acute and convalescent HFRS. As sgp130 can inhibit IL-6 trans-signaling,^19^ a decrease in sgp130 may have pathogenic consequences. In support of this notion, decreased sgp130 levels have been described in patients with type 2-diabetes,^44^ and in patients with coronary artery disease.^45–47^ Furthermore, increased sgp130 levels have been associated with decreased odds of myocardial infarction.^48^ The inverse correlation found between plasma sgp130 and number of interventions during acute HFRS may indicate that patients with low sgp130 levels also have more severe symptoms. Albumin has been suggested as a marker of vascular permeability.^49^ Thus, the positive correlation between sgp130 levels and serum albumin levels during acute HFRS may suggest a link between low sgp130 levels and increased vascular leakage. Interestingly, in patients with coronavirus disease 2019 (COVID-19), low levels of serum albumin have been associated with more severe pulmonary symptoms.^50^ Analyses of IL-6 receptor levels and of possible correlations to risk for severe and fatal outcome in HPS patients could help decipher the role of IL-6 trans-signaling in hantavirus-induced pathogenesis, as these patients in general are more severely ill than HFRS patients.

The skewed IL-6-receptor balance during acute HFRS, as indicated by the increased sIL-6R/sgp130 ratio, suggests an increased potential of IL-6-trans-signaling in patients. Importantly, while classical signaling via membrane-bound IL-6R mainly seems to have homeostatic, protective, effects, trans-signaling has recently emerged as the driver of IL-6 mediated pathogenesis.^18^ In most HFRS patients, plasma IL-6 levels are not as strongly elevated as observed in HPS patients. Given the capacity of endothelial cells to produce large amounts of IL-6, it is possible that the concentration of IL-6 is higher at local sites of infection than systemically. It is further possible that highly vascularized sites, such as the lungs, have very high local concentrations of infected endothelial cell-derived IL-6, and as a consequence are more exposed to IL-6 trans-signaling. This scenario is consistent with the observations that lungs are heavily affected during HPS, and that almost all lung endothelial cells are infected in some patients.^51–53^ Altogether, our data suggest altered concentrations of sIL-6R and sgp130 during hantavirus-infection, which may increase the likelihood of IL-6 trans-signaling in infected endothelial cells and increased vascular permeability as a consequence.

Treatment strategies targeting IL-6 signaling have proven successful in for example rheumatoid arthritis and for treatment of COVID-19.^18,54,55^ Further studies investigating the role of IL-6 in the pathogenesis of HFRS and HPS will be valuable for the assessment of the relevance of IL-6-targeting therapeutics in severe hantavirus infections. In conclusion, we show that PUUV-infected endothelial cells produce IL-6 that, in an IL-6 trans-signaling dependent manner, strongly affect infected endothelial cell functions. We further show a correlation between IL-6 trans-signaling potential and severity in HFRS patients. These findings suggest that IL-6 trans-signaling may represent a treatable target in HPS and severe HFRS.

## Data Availability

All data produced in the present work are contained in the manuscript

## Data sharing

All data are available in the main text or the supplementary materials.

## Acknowledgements

We thank the patients and volunteers who contributed with clinical material to this study. C. Ahlm received fundings from Region Västerbotten and Umeå University, project numbers RV-579011, RV-734361, and RV-965866, and from the Heart-Lung Foundation, project numbers 20170334 and 20150752. J. Klingström received fundings from the Swedish Research Council, project number 2018-02646, and from the center for medical innovation (CIMED), project number 2020-0141.

The funders played no role in the design of the study, data collection, data analysis, interpretation of results or writing of the paper.

## Author contributions

Conceptualization: KTM, JK

Methodology: KTM, LN, WC, JK

Investigation: KTM, LN, WC, TT, CA

Analyzed data: KTM

Validated data: KTM, LN

Visualization: KTM

Funding acquisition: CA, JK

Project Administration: KTM, JK

Resources: JWB, TT, CA

Supervision: KTM, JK

Writing – original draft: KTM, JK

Writing – review & editing: KTM, LN, WC, JWB, TT, CA, JK

