## Supplementary figures 1 and 2 for "IL-6 trans-signaling mediates cytokine secretion and barrier dysfunction in hantavirus infected cells and correlate to severity in HFRS"

**Supplementary material**

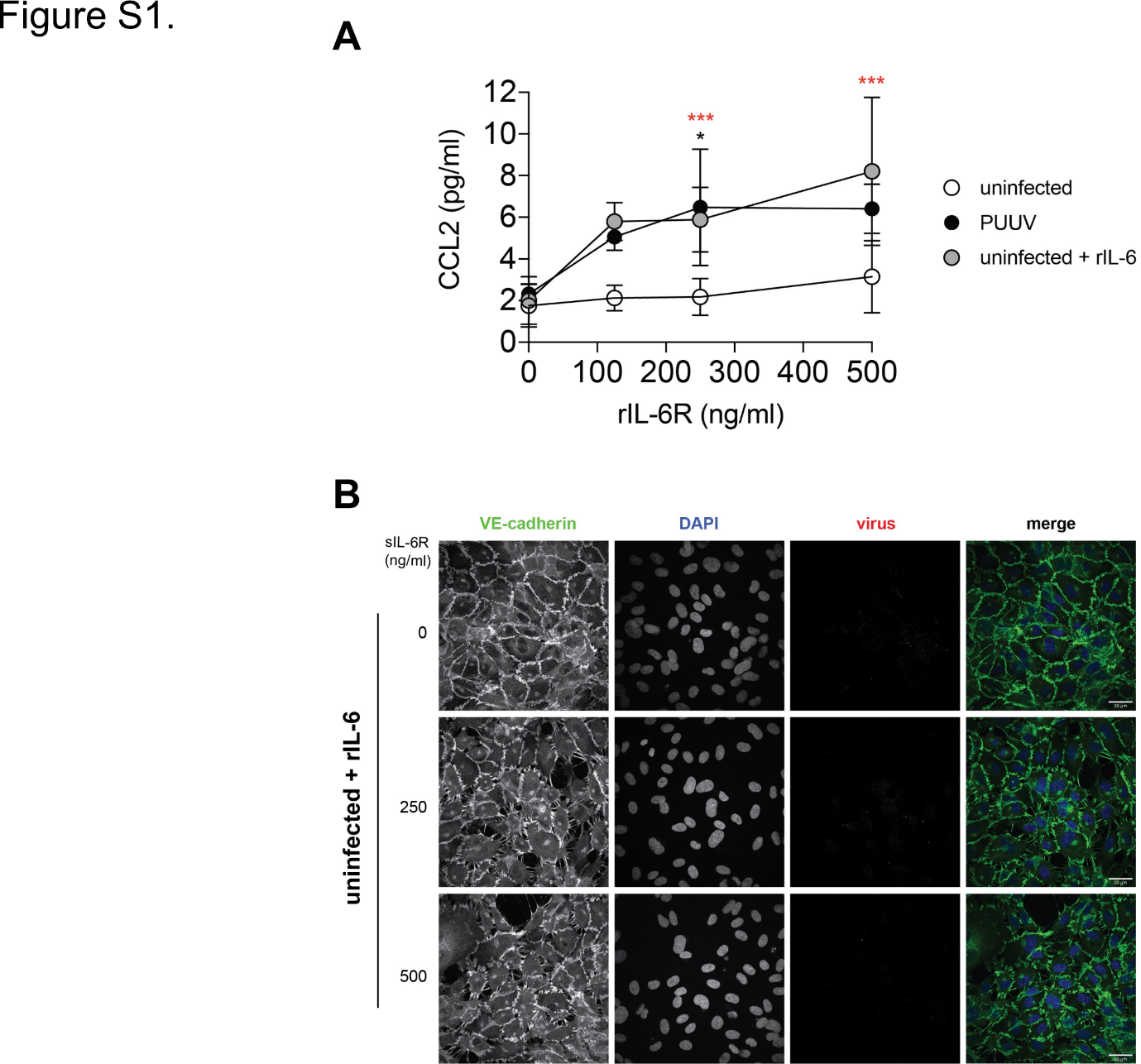

**Figure S1.** **Treatment with rIL-6 and sIL-6R in uninfected endothelial cells causes increased CCL2 secretion and affects the endothelial cell barrier.** (**A**) Levels of CCL2 in supernatants of uninfected and infected HUVECs treated with sIL-6R and/or rIL-6, or left untreated (n=3, except n=2 for 125 ng/ml). (**B**) Immunofluorescence images showing expression of DAPI (blue), virus (red), and VE-cadherin (green). Representative images of three independent experiments are shown. Symbols depict mean and error bars indicate SD. Two-way ANOVA followed by Dunnet's or Šídák's multiple comparison test. Black asterisks indicate significance when comparing PUUV to uninfected. Red asterisks indicate significance when comparing each sIL-6R-treated conditions of PUUV-infected cells with untreated PUUV-infected cells. *, p<0·05; **, p<0·01; ***, p<0·001, ****, p<0·0001.

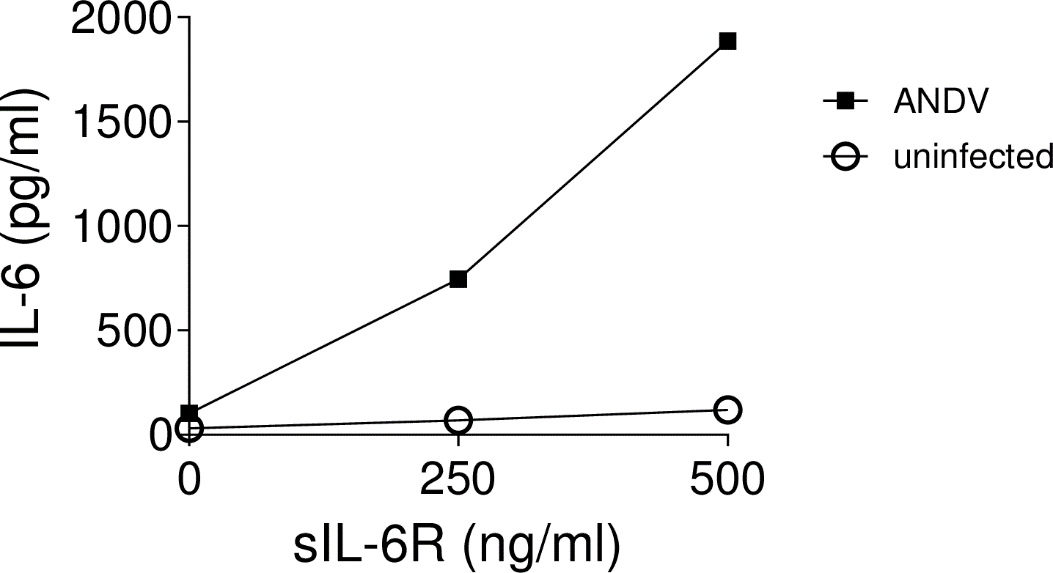

**Figure S2. rIL-6 treatment of ANDV-infected endothelial cells causes increased IL-6 secretion.** Levels of IL-6 in supernatants of uninfected and ANDV-infected HUVECs treated with sIL-6R or left untreated (n=2; data shown represent the mean value).
